# GPS-Health: A Novel Analytic Infrastructure for Capturing, Visualizing, and Analyzing Multi-Level, Multi-Domain Geographically Distributed Social Determinants of Health

**DOI:** 10.1101/2025.01.03.25319962

**Authors:** Shuo Jim Huang, Esa M. Davis, Thu T. Nguyen, Justin R. Brooks, Olohitare Abaku, Se Woon Chun, Oluwadamilola Akintoye, Sinan Aktay, Matthew Chin, Matthew Bandos, Sunil Pateel, Vineeth Gohimukkula, Victor Felix, Anup A. Mahurkar, Rozalina G. McCoy

## Abstract

**Background:** Health disparities across a range of conditions and outcomes exist across the life course and are driven by the uneven geographic distribution of multidimensional social determinants of health (SDOH). Previous multidimensional measures of SDOH (e.g. Area Deprivation Index, Social Vulnerability Index, Social Deprivation Index) collapse multiple measures into a single summary value applied to everyone living within a predefined map unit, engendering construct and internal validity issues.

**Methods:** We present a new SDOH data approach: the Geographic Patterns of Social Determinants of Health (GPS-Health). We use a theoretical framework weaving together kyriarchy, intersectionality, and structural violence to select SDOH domains that can elucidate how individuals experience multidimensional spatial distributions of SDOH. We apply the approach to Maryland.

**Results:** Our dataset includes 2,369,365 property parcels, from which we calculate distances to 8 types of SDOH exact locations.

**Discussion:** GPS-Health will aid in the understanding of how the SDOH influence individual health outcomes.

## Introduction

Health disparities across a range of conditions and outcomes exist across the life course^1,2^. Inequities in the social determinants of health (SDOH) are a key driver of health disparities^3–9^. SDOH refer to the conditions of daily life^10^ including housing, wealth and economic security, healthcare access, food access, education, safety, civic access, and pollution and environmental hazards^5,11^. SDOH are estimated to influence between 30% and 70%^12,13^ of health outcomes (depending on included domains). Therefore, the uneven geographic and spatial distribution of resources and hazards in each individual’s social and physical environment^14,15^ constitutes a critical multidimensional SDOH context that impacts their health.

Historically, SDOH factors have been examined in health-related research as mostly unidimensional constructs^5^ (such as income). Developments in the last two decades of geographic area indices of SDOH—such as Area Deprivation Index^15–17^ (ADI), Social Vulnerability Index^18^ (SVI), and Social Deprivation Index^19^ (SDI)—sought to illustrate the importance of the multidimensional and contextual nature of SDOH within defined geographic areas. While a major advancement over unidimensional constructs, these area indices rely on methodological choices that limit their ability to capture the multidimensional nature of SDOH factors as it applies to individuals. These methodological choices include: 1) using convenient map units as the unit of analysis (such as census tracts or counties), and 2) using a summary (or composite) index value across a range of different measures^20^.

These two intertwined methodological choices (summarizing multidimensional SDOH factors into a single score and applying that score to a standard map unit) engender the following internal and construct validity issues: 1) By collapsing a multidimensional SDOH context into a single summary measure, area indices cannot distinguish between different SDOH permutations^20^ (e.g. poor housing conditions vs. poor food access), particularly if different SDOH have variable effects on different people and communities. 2) By collapsing and then applying a single geographic measure across an entire map unit to every individual living within that map unit, area indices mask the heterogeneity of proximity-based exposures across that map unit^21,22^. Thus, area indices of SDOH may not reflect the true multidimensional SDOH context that each observed individual lives in^21,23^.

A new SDOH data approach is therefore needed to address these validity issues, one enabled by advances in robust computational methods and the availability of granular level data that capture the wide range of SDOH. This approach would identify not only what factors are included in a multidimensional SDOH model, but also how those factors are displayed, measured, and centered on an appropriate unit of analysis to better explain health disparities. The purpose of this paper is to present a novel theoretical framework and methods for such an approach. To demonstrate our method, we build an initial version of a dataset, which we term GPS-Health (**G**eographic **P**atterns of **S**ocial Determinants of **Health**) using data from Maryland.

### Theoretical framework

The importance of power and social structure to health outcomes^4^ is explicitly mentioned in our most commonly used definitions of health disparities^6,24^, tying them explicitly to social disadvantage^25^. Yet common usage of SDOH in spatial research on clinical outcomes and interventions has often disconnected this analysis of power, social structure, and hierarchy from SDOH distribution^26^.

To inform our work, we employ three theories on how power influences social structure: kyriarchy, intersectionality, and typologies of violence. **Figure 1** illustrates how each is used in our conceptual framework. Our conceptual framework shows how societal power structures influence land use policies that in turn generate the geospatial distribution of SDOH.

**Figure 1.**
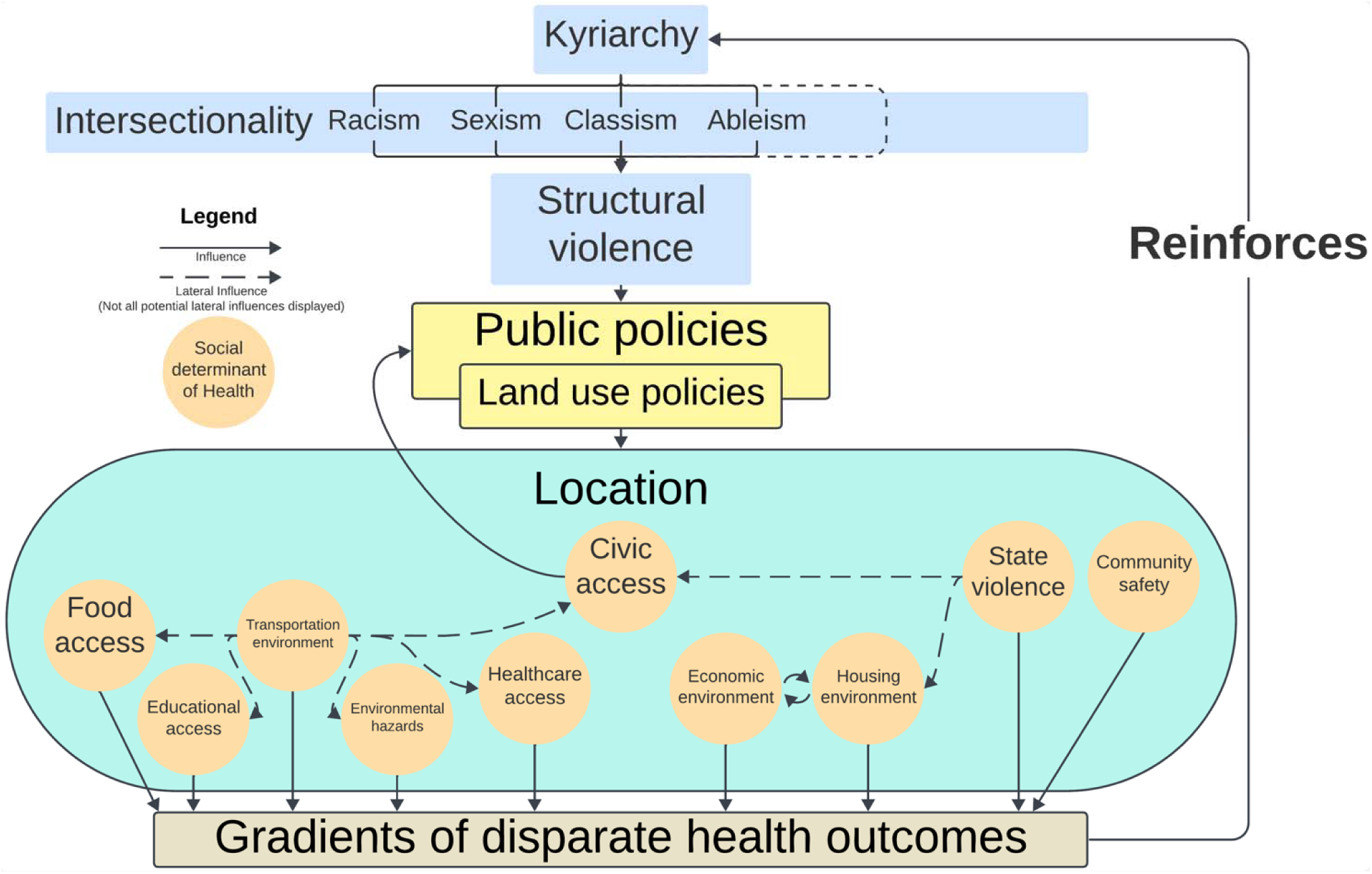
Conceptual Framework. Lateral influence arrows do not show all possible influences among social determinants of health domains. Short dotted line to the right of “Ableism” is for other types of Kyriarchical social domination that are not shown here.

Fiorenza’s kyriarchy^27^ posits that oppressive social systems are interconnected structures of social domination. Under this framework, racism and sexism—for example—are not separate systems from each other, but rather interlocking and mutually reinforcing systems resulting in particular forms of social hierarchy along race and gender. As other authors have noted^28^, this “dovetails” with Crenshaw’s intersectionality framework^29^, which posits that those whose identities sit at the intersections of multiple historically marginalized identities (for example Black women by race and gender^29^) do not face just the additive oppressive burdens imposed by society on each identity, but rather a different, often multiplicative one.

These two frameworks are already in use for studies of how people experience spatial vulnerability in academic fields such as Planning^28,30^, which studies land-use policies. However, while these theories help illuminate how and why spatial vulnerability exists, they do not describe the mechanisms by which the spatial distribution of SDOH occurs. For that, we employ Galtung’s typologies of violence^31^, specifically structural violence^31,32^ defined as the systematic deprivation of the conditions of life (e.g. policies preventing a community from accessing food^32^).

Structural violence’s deprivation of the necessary conditions of life^31^ thus fits neatly within commonly accepted definitions of SDOH as “conditions of daily life” ^10^. By doing so, structural violence posits a clear mechanism to the distribution of SDOH: societal structures shape the building of an individual’s lived environment^14,33–37^—via processes and policies like land use zoning and permitting^33,34^—to impose strongly patterned selective access to resources and exposures to hazards^38–40^. This selective access in turn impacts healthcare delivery and health outcomes^35–37,41–55^. Kyriarchy explains that structural violence is directed toward individuals who have been historically marginalized and deprived of political power, for example by race for Black and Indigenous people^28^. Intersectionality explains how that violence compounds, which in turn explains the lived experiences of individuals marginalized by overlapping forms of structural violence^29,30^. Thus, we utilize our combined framework to build a model of the geospatial distribution of SDOH that connects oppressive and persistent social structures of advantage and disadvantage to the multidimensional confluences of SDOH resources and hazards that surround individuals based on where they live.

### Selection of SDOH thematic domains

Using our theorized pathway that structural violence shapes spatial access to SDOH, we selected SDOH thematic domains that explain the geospatial distribution of SDOH, with detailed explanations for each in **Table 1**. Our thematic domains include housing environment, economic environment, healthcare access, food access, educational access, transportation environment, civic access, community safety, state violence, and environmental hazards. While not mutually exclusive (for example, state violence can deprive people of secure housing^56^), these domains provide a starting point to select datasets and pair them with existing models of SDOH.

**Table 1:** Selection of thematic domains.

| **SDOH domain** | **Explanation of how structural violence impacts distribution of domain** |
| --- | --- |
| Housing environment | Control over housing access and quality has historically been an aspect of structural violence<sup>14,33,56,76–83</sup>. Housing is an essential necessity for health and life and its quality or lack of is strongly associated with health, social, and economic outcomes<sup>14,36,37</sup>. |
| Economic environment | Economic positioning and access to economic activity has been used to determine power hierarchies<sup>84–86</sup>. In addition to its use as shelter, housing is also both a major store of potential household wealth<sup>33,35,87</sup> and conversely a potential site of cost burden and exposure to extractive rent-seeking practices<sup>82,86</sup>. Neighboring land use and economic development is also an aspect of economic well-being<sup>88,89</sup>. |
| Healthcare access | Systematic and targeted deprivation of healthcare access is used as an element of social control and power<sup>90–92</sup>, including at the macro level with restrictions on sites providing abortion and other aspects of reproductive care<sup>91,93</sup>, restrictions on sites providing tailored care for gender and sexual minorities<sup>94–96</sup>, and strong geographic racial, political, and rural patterning on access to insurance<sup>41,97</sup>. These can manifest as systematic racial and class disparities in geographic access to quality healthcare<sup>98</sup>. |
| Food access | Food access and famine are longstanding methods of structural violence<sup>99,100</sup>. Restrictions on food and nutrition have been used as weapons of war<sup>32,101,102</sup> as well as ways to control politics and populations<sup>32,103</sup>. More recently, food “deserts” have been used as evidence of food apartheid and lack of democratic responsiveness<sup>104–108</sup>. |
| Educational access | While educational attainment alone is not fully protective against structural violence and discrimination<sup>109</sup>, disparate availability of educational resources and quality has been used historically to enshrine intergenerational patterns of structural violence<sup>79,110,111</sup>. This has included both historical de jure school segregation (including in Maryland) as well as continuing de facto attempts to continue school segregation<sup>79,111</sup>, including through housing policy<sup>79</sup>. |
| Transportation environment | Restrictions on or differential facilitation of movement has long been used as a method of political control<sup>90</sup> and structural violence<sup>112</sup>. Those with lower access to transportation and transit have greater claims on their time and resources<sup>113</sup>, are less able to access health<sup>113–116</sup>, civic<sup>117</sup>, and social infrastructures<sup>113,118,119</sup>, and are less able to avoid environmental hazards<sup>120</sup>. At the same time, transportation infrastructure building has often been racially and/or economically exclusionary<sup>121</sup>, or has been used to disrupt housing and neighborhood networks<sup>122</sup>. Transportation infrastructure can be a source of environmental pollution<sup>123</sup>. Transportation modalities can potentially be hazardous, particularly in the form of vehicle crashes<sup>124</sup>. |
| Civic access and democratic responsiveness | By our theoretical framework, civic access is ultimately one of the more direct measures of direct retention of control over power. Attempts to directly disenfranchise voters by geographic racial distribution are well-documented<sup>125</sup>. Civic access influences democratic responsiveness and policy changes around the distribution of SDOH and public resources that impact health<sup>41,126,127</sup>. |
| Community safety | Rather than fully stochastic processes, community violence follows patterns of political repression<sup>128–130</sup>. Seemingly stochastic community violence against people from intersecting historically marginalized groups, including against women<sup>131</sup>, Black, Indigenous, Latino, Asian, and other people of color<sup>128,131</sup>, |
|  | LGBTQIA+<sup>131,132</sup>, and religious minorities<sup>133,134</sup> have been a hallmark of political violence, as well as at times deliberately encouraged by political leaders<sup>133,134</sup>. Geographically uneven policy choices around the supply of weapons may also be a factor in disparate community violence<sup>135</sup>. |
| State violence | Direct state use of violence and coercion often undergird structural violence<sup>32</sup>. State violence, including negative police interactions<sup>129,132,136–138</sup>, police killings<sup>139,140</sup>, incarceration<sup>90,141,142</sup>, and evictions<sup>82,143</sup> are well documented in their deleterious effects on health, educational, and economic outcomes<sup>92,141,144–146</sup>. Additionally, selective state violence has been used as an arbiter of “functional citizenship” where police violence functionally demarcates the boundaries of full state protection and civic participation<sup>136</sup>. Direct state violence is also used to enforce other forms of structural violence. For example, the use of state direct violence to enforce evictions<sup>56,83</sup> and control the movements of people experiencing homelessness<sup>147</sup> reinforces the differential distribution of housing as a SDOH. |
| Environmental hazards | While residential exposures to some environmental hazards are more obviously manmade, such as pollution<sup>30,148–150</sup>, the differential siting of housing and disaster mitigatory infrastructure also exposes people from different social strata to differential risk of “natural” hazards<sup>151</sup>, including some highly publicized examples in the USA in the last few decades from wildfires<sup>152,153</sup> and flooding events<sup>154,155</sup>. |

SDOH: Social determinants of health.

In this iteration of GPS-Health, we excluded domains that are primarily employment and labor related (such as site-specific labor violations or discriminatory hiring practices). While labor relations and conditions are a key component of power relations, potential exploitation, and structural violence in society^57,58^, we do not yet have a way to assess the particular employment features of where individuals work.

### Implementation of GPS-Health in Maryland

The state of Maryland is a mid-sized coastal state in the USA. It is racially diverse with higher population proportions of Black/African American and Asian people than the national average^59^. Maryland also has both major urban and rural areas, counties in both the top 99^th^ percentile and bottom 25^th^ percentile of median income in the USA^60^, and highly disparate outcomes^61–65^. It is historically important, being an Union state with de jure slavery during the Civil War^66^, and anti-Black Jim Crow laws in the postbellum period^46,66^. Finally, Maryland implements a global hospital budget Medicare waiver^67^ and collects high quality health outcomes data through multiple state-wide institutions^68^. In short, Maryland is a microcosm of the broader USA, with enough variation and size to be able to tease out differences in SDOH profiles.

### Objective

Our objective is to define a method for layering contextual data of social determinants of health for individual residences that can be used in multi-dimensional analyses, using data from Maryland.

## Methods

### IRB statement

The University of Maryland Baltimore Institutional Review Board determined this Not Human Subjects Research.

### Datasets curation

We merged multiple datasets (**Table 2**) to establish sites of SDOH hazards and resources from each of the following domains: housing environment, economic environment, healthcare access, food access, educational access, transportation environment, civic access, community safety, state violence, and environmental hazards. Our selected datasets cover all theorized domains for Maryland.

**Table 2:**
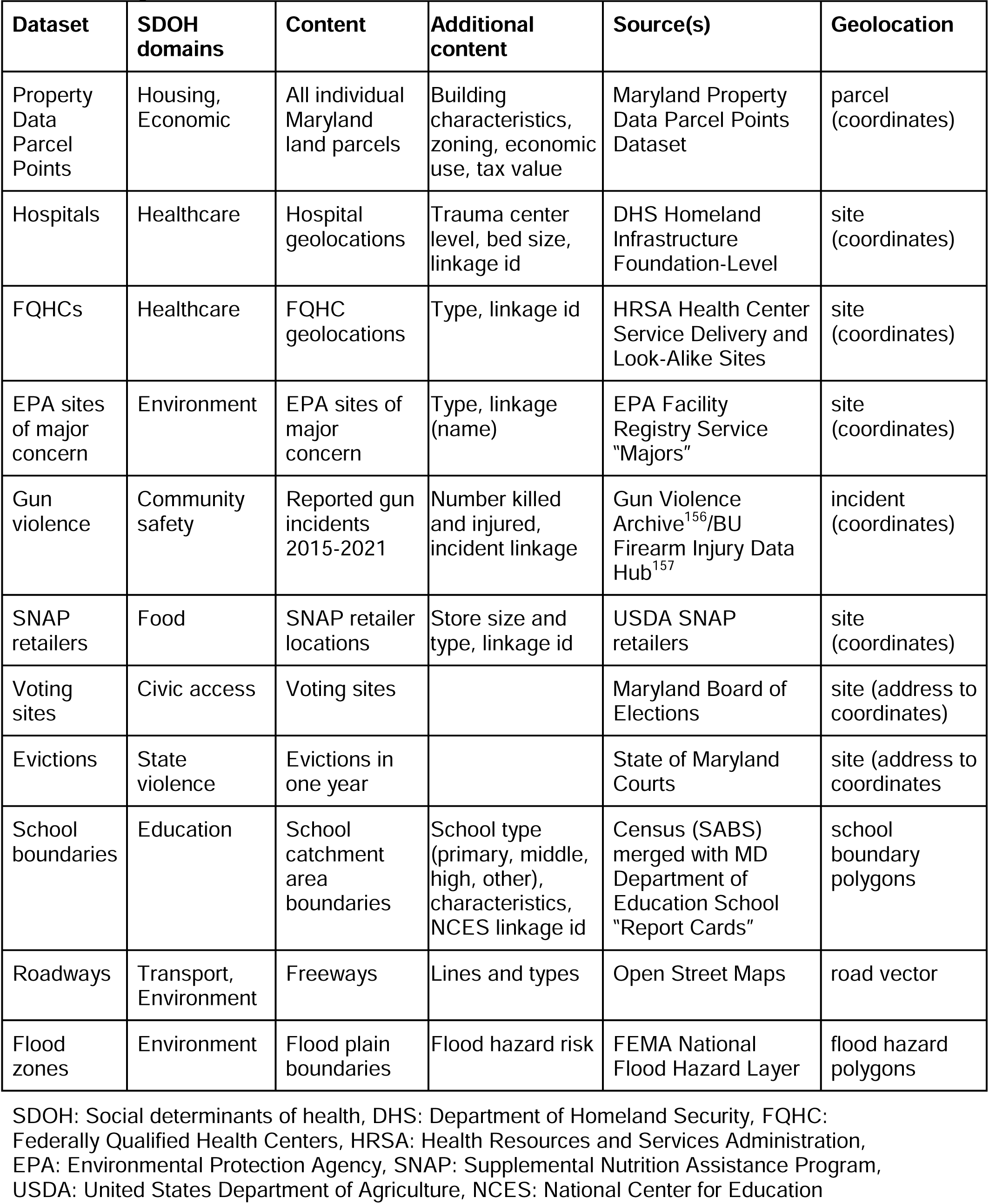
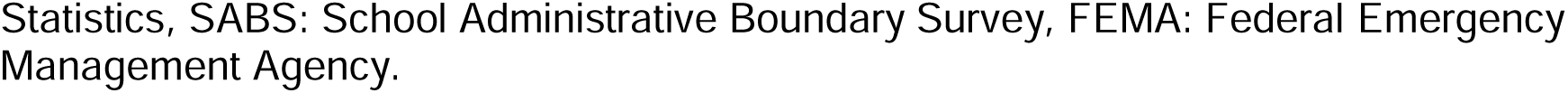
Merged dataset.

| **Dataset** | **SDOH domains** | **Content** | **Additional content** | **Source(s)** | **Geolocation** |
| --- | --- | --- | --- | --- | --- |
| Property Data Parcel Points | Housing, Economic | All individual Maryland land parcels | Building characteristics, zoning, economic use, tax value | Maryland Property Data Parcel Points Dataset | parcel (coordinates) |
| Hospitals | Healthcare | Hospital geolocations | Trauma center level, bed size, linkage id | DHS Homeland Infrastructure Foundation-Level | site (coordinates) |
| FQHCs | Healthcare | FQHC geolocations | Type, linkage id | HRSA Health Center Service Delivery and Look-Alike Sites | site (coordinates) |
| EPA sites of major concern | Environment | EPA sites of major concern | Type, linkage (name) | EPA Facility Registry Service “Majors” | site (coordinates) |
| Gun violence | Community safety | Reported gun incidents 2015-2021 | Number killed and injured, incident linkage | Gun Violence Archive<sup>156</sup>/BU Firearm Injury Data Hub<sup>157</sup> | incident (coordinates) |
| SNAP retailers | Food | SNAP retailer locations | Store size and type, linkage id | USDA SNAP retailers | site (coordinates) |
| Voting sites | Civic access | Voting sites |  | Maryland Board of Elections | site (address to coordinates) |
| Evictions | State violence | Evictions in one year |  | State of Maryland Courts | site (address to coordinates) |
| School boundaries | Education | School catchment area boundaries | School type (primary, middle, high, other), characteristics, NCES linkage id | Census (SABS) merged with MD Department of Education School “Report Cards” | school boundary polygons |
| Roadways | Transport, Environment | Freeways | Lines and types | Open Street Maps | road vector |
| Flood zones | Environment | Flood plain boundaries | Flood hazard risk | FEMA National Flood Hazard Layer | flood hazard polygons |

SDOH: Social determinants of health, DHS: Department of Homeland Security, FQHC: Federally Qualified Health Centers, HRSA: Health Resources and Services Administration, EPA: Environmental Protection Agency, SNAP: Supplemental Nutrition Assistance Program, USDA: United States Department of Agriculture, NCES: National Center for Education
Statistics, SABS: School Administrative Boundary Survey, FEMA: Federal Emergency Management Agency.

We linked and merged these datasets based on geolocation. All included datasets contain at least one of the following elements that can establish geographic location: geocoordinates (e.g. latitude/longitude), geolocated lines or polygons for features that span between locations (such as a road) or cover a larger area (such as a flood zone), or street address. Thus—using our merged dataset—we could calculate the distance from each geolocated property parcel to a discrete geolocated SDOH resource or hazard, or that parcel’s geographic location within a larger SDOH resource or hazard’s boundaries. These distance measures and colocations within specific boundaries served as the core of our example analytic dataset.

If a dataset uses street address for location, we used an address cleaning algorithm (removing non-standardized address features such as suite numbers) followed by an OpenCage geocoder^69^ to assign latitude/longitude geocoordinates to the address^70^. From this geocoding step, we retained sites that have high bounding box confidence (< 0.5 km) and that return an actual address or address-like feature^70^. As much as possible, we used datasets that already contained geocoordinates to bypass the added geocoding step.

Several of our geolocated datasets include administrative identifier variables, which could be further extended to merge in additional layers of contextual data from sources that lack built-in location data. We merged in Maryland state Department of Education school data to our school catchment area boundaries using such an administrative linkage as an example.

Since some geospatial exposures and resources do not necessarily follow state lines (e.g. pollution sites or healthcare facilities), we drew data on site locations from the national level— where available—including additional sites outside of Maryland to better cover potential exposures. This has the added benefit of facilitating the building of GPS-Health datasets in other states in the future.

The individual residence dataset at the core of our data infrastructure is the Maryland Property Data Parcel Points of all Maryland land parcels^71^. Land parcels are the individual, legal real estate property divisions, upon which residential and commercial structures may be built. This dataset includes parcel centroid latitude and longitude coordinates, from which we can establish the geolocation of all Maryland residences to a high degree of specificity. The dataset also includes parcel level data, including assessed tax value, built structures, and land use zoning category, amongst others. This data is publicly available, current to the month, and free to use. Please see **Table 2** for all the other datasets we used, which are all similarly publicly available and free to use.

### Analysis

We characterized the dataset in our results section. Additionally, using QGIS 3.34.2, we performed nearest neighbor great circle arc (“as the crow flies”) distance calculations on distance to single location exposures, and reported summary statistics of those distance calculations in degrees.

## Results

We reported counts of SDOH hazards and resources in **Table 3**. There are 2,369,365 property parcels in the state of Maryland. We found 8,013 hospitals (73 in Maryland) and 1770 FQHCs nationwide (161 in Maryland). Our dataset included 227,292 gun violence incidents (6923 in Maryland) from 2015 to 2021. There were 16,961 Environmental Protection Agency (EPA) sites of major concern (2,273 in Maryland). Nationally, there were 262,338 Supplemental Nutrition Assistance Program (SNAP) retailers, (3794 in Maryland). Maryland had 14,111 evictions in one year, and 2,153 voting sites. Maryland contained 9,784 freeway road segments. There were 1,286 non-tertiary schools of all types in Maryland, and 37,094 flood hazard zones.

**Table 3:** Results.

|  | count<br>(Maryland) | count<br>(USA/other<br>states) | mean<br>distance | std dev<br>distance |
| --- | --- | --- | --- | --- |
| Property<br>Data Parcel<br>Points | 2369365 |  |  |  |
| Hospitals | 73 | 8013 | .0698 | .0587 |
| FQHCs | 161 | 1770 | .0740 | .0647 |
| EPA sites of<br>major<br>concern | 2273 | 16961 | .0200 | .0206 |
| Gun violence | 6923 | 227292 | .0212 | .0280 |
| SNAP<br>retailers | 3794 | 262338 | .0141 | .0161 |
| Voting sites | 2153 |  | .0183 | .0209 |
| Evictions | 14111 |  | .0193 | .0273 |
| Freeway<br>segments | 9784 | 31386 <sup>#</sup> | .0725 | .1185 |
| Flood zones | 37094 |  |  |  |
| School<br>boundaries | 1286 |  |  |  |

Mean and standard deviation of great arc distances reported in degrees. FQHC: Federally Qualified Health Centers, EPA: Environmental Protection Agency, SNAP: Supplemental Nutrition Assistance Program. <sup>#</sup> Additional freeway segments were included from border regions of states surrounding Maryland.

Mean and standard deviation of great arc distance calculations between all individual parcels and the nearest neighboring exact location SDOH hazard and resource in each domain were also reported in degrees in **Table 3**. Mean exact location SDOH distances ranged from a low of .0141 degrees to a SNAP food retailer site to a high of .0740 degrees to a health center site. Standard deviations of exact location SDOH distances ranged from a low of .0161 degrees to a SNAP retailer site to a high of .1185 degrees to a freeway.

Parcel-level distributions of distance measurements for each single location sites are shown via histograms in **Figure 2**. Distance measurements are all right-skewed. There were particularly strong peaks at the left tail of the distributions (less than .01 degrees) for proximity to a gun violence incident and to an eviction, while there were more flattened distributions for proximity to hospitals and FQHCs.

**Figure 2:**
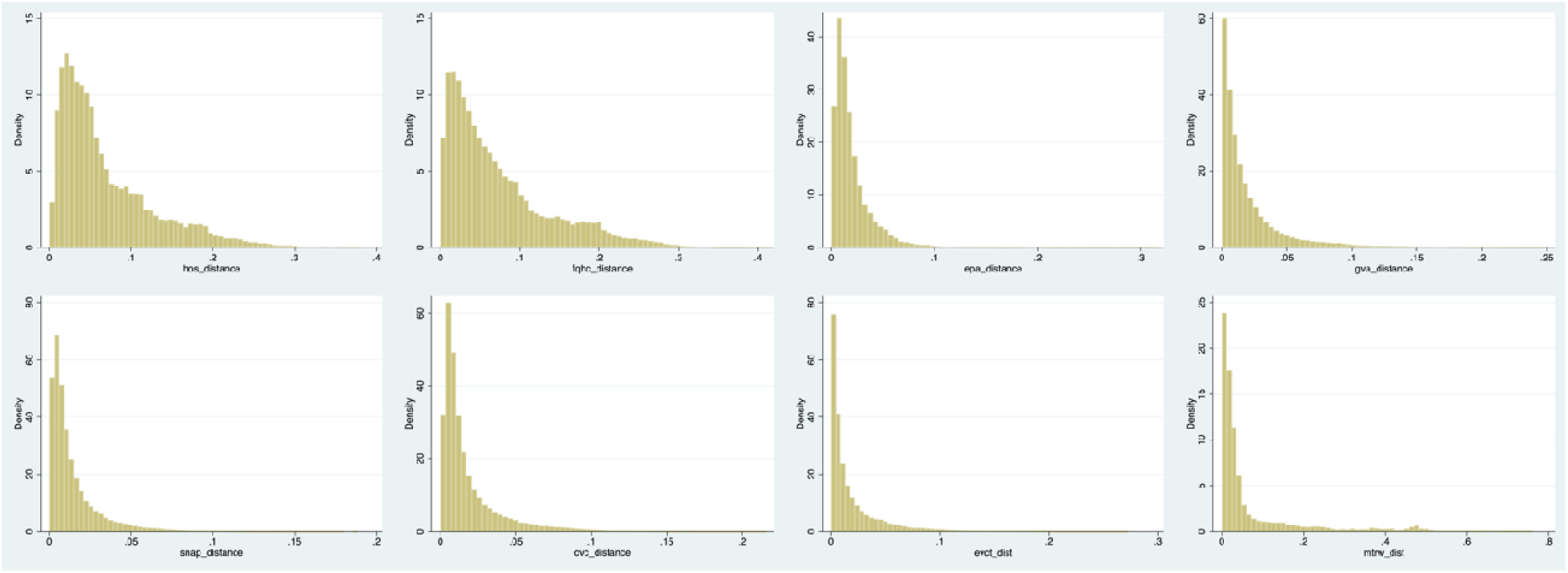
Distributions of parcel distances to exact location SDOH hazards and resources. Counter-clockwise from upper right: gun violence incidents, Environmental Protection Agency (EPA) major registered facility sites, Federally Qualified Health Centers (FQHCs), hospitals, Supplemental Nutrition Assistance Program (SNAP) retailers, voting locations, eviction sites, freeways (access-controlled highways)

We displayed all SDOH resources and hazards exact site locations and freeway roads in **Figure 3**. Sites tended to cluster in and around major metropolitan centers and along major transportation lines, with decreasing density in less populated areas.

**Figure 3:**
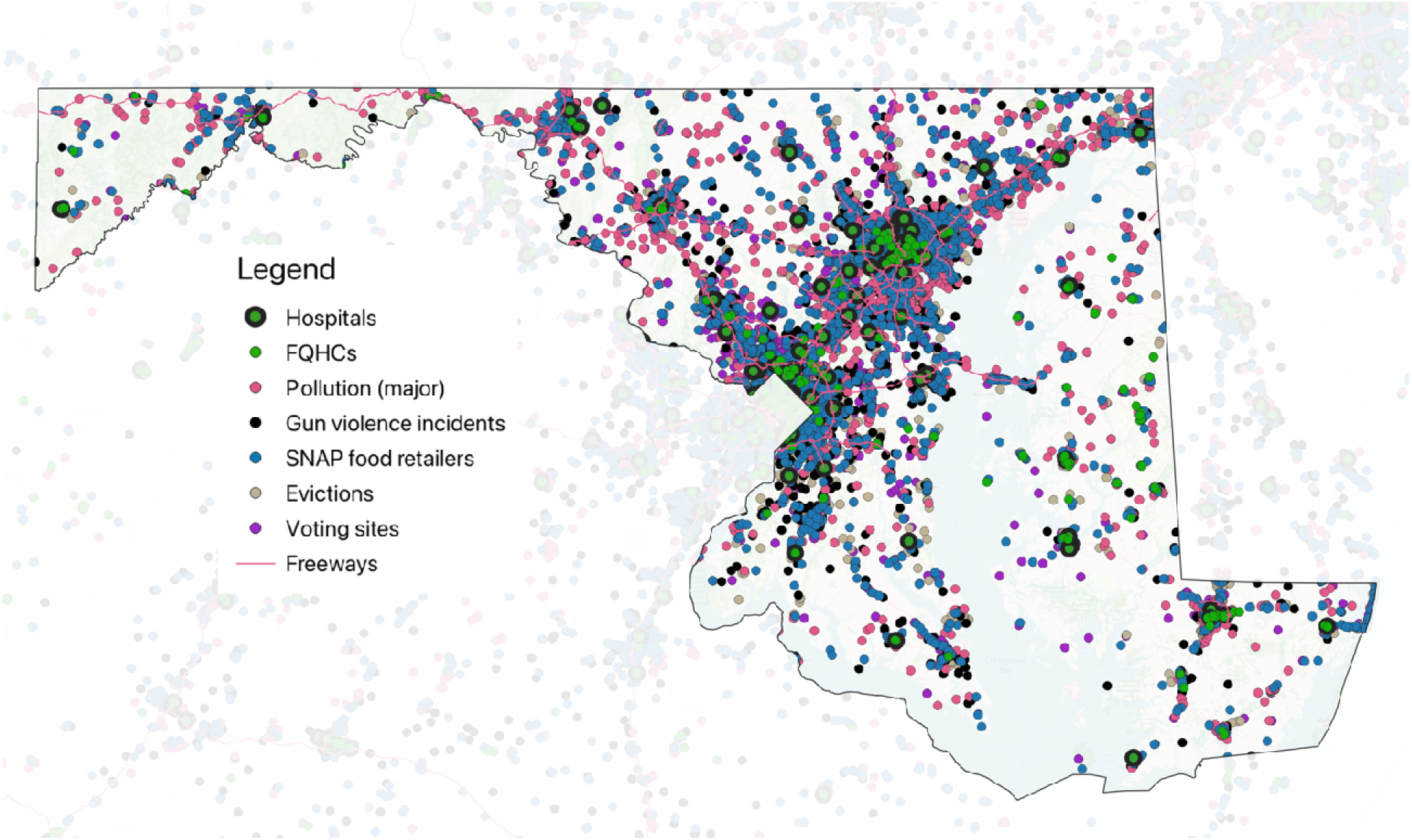
All individual location Social Determinants of Health resources and hazards in Maryland. FQHC: Federally Qualified Health Centers, SNAP: Supplemental Nutrition Assistance Program. Basemap layer is of Maryland (boundary outlined) from Open Street Maps. Faded areas outside of Maryland’s boundaries are from surrounding states.

The de-identified analytic dataset is available in a repository at: https://doi.org/10.5281/zenodo.14422743. The dataset includes distances to the nearest neighboring SDOH resource and hazard in each subdomain along with additional contextual data elements and available linkage elements.

## Discussion

We presented a theoretical framework, method, and summary statistics for GPS-Health, a data approach for use in analysis of multidimensional residential SDOH, which we applied to the state of Maryland. Our theoretical framework combines the framework of structural violence with those of kyriarchy and intersectionality to guide the selection of our data domains and can be enriched and updated in real-time with additional data. GPS-Health can provide individual household level, rather than map unit area level SDOH measurements, which advances the field of SDOH research by allowing for the capture and analysis of multi-dimensional, intersectional, and individually contextualized SDOH constructs. The method we employed addresses internal and construct validity issues of using map unit indices for individual outcomes, reducing heterogeneity loss from not collapsing measures to a map unit, and allowing for future data-driven clustering of SDOH at the address level.

GPS-Health currently includes specific location data for hazards and resources important for each of our theorized domains: housing environment, economic environment, healthcare access, food access, educational access, transportation environment, civic access, community safety, state violence, and environmental hazards. Several of these SDOH domains (healthcare access, food access, civic access, community safety, state violence, and environmental hazards) go beyond those captured by dimensional reduction methods in ADI, SVI, and SDI^15–19^. Even in domains shared with ADI, SVI, or SDI, since we used individual environs rather than a map area, GPS-Health measures different constructs^23^ while maintaining the ability to study wider phenomenon at a greater resolution^21^. Finally, since our framework theorizes that structural violence drives differences in SDOH distribution for different social groups as the mechanism to uphold social domination, we do not directly incorporate demographics (such as race/ethnicity, English language proficiency, or age) into our measures. This allows GPS-Health to be used in analysis of *how* disparities in SDOH distribution drive health disparities. Combined, these features may enable the type of multidimensional, individual-level analysis that would allow clinicians to tailor specific referrals to supportive services to their patients and communities to understand the types of health risks faced by different segments of their populations.

Although built exclusively using publicly available data, our dataset contains granular data on individual residences, which should be treated with similar safeguards as those afforded to protected health information. Despite being available and accessible in the public domain, these data may nevertheless be considered private by individuals depending on context^72^. For this reason, our project was reviewed by our institution’s IRB and the publication includes only deidentified data. Our planned use of these data will be within a secure research environment to prevent disclosure and loss of confidentiality. Moreover, the broader societal implications of the use of such granular data need to be considered, particularly for populations historically marginalized by societal power structures. Granular, broad-scoped data should only be used in accordance with principles of beneficence^73^ and in accordance with regulations for the protection of human subjects. A major concern is the use of these publicly available data for “algorithmic redlining”^74,75^, such as restrictions on service provision in healthcare, social services, housing, and other programs. Given the widespread existence of data brokers and the relative ease in which these types of data can be combined, legal policies need to be put in place to protect against data use for discriminatory purposes.

### Limitations

Given this is an initial iteration of our dataset, it has a few limitations, as each domain of SDOH measurement likely contains other resources and/or hazards. Our infrastructure and method, however, will allow the future inclusion of additional layers of contextual data.

Data completeness also relies on the completeness of source data. We used data sources that are either widely used already for research purposes or are otherwise administrative datasets used commonly for planning of land use (such as the property parcel dataset).

The scope of our dataset is focused on Maryland. However, since Maryland is a unique state due to its highly heterogeneous population, we anticipate our methods piloted in Maryland will be applicable to other areas of the USA.

Since our data relies on property parcels, we cannot adequately capture SDOH that are not related to the built environment. One clear exclusion is labor-related SDOH as mentioned before, but this is an area where additional data can be added as it becomes available in the future. We are also unable to capture, for example, gender dynamics within a household or the residential experiences of people experiencing homelessness. Future work should look to new methods and frameworks to be able to combine and layer these aspects of SDOH as well.

## Conclusion

By layering together data on each individual’s experience of the geographic distribution of SDOH, GPS-Health is a paradigm shift in how SDOH are studied and used to understand and improve health and health disparities. Future work could apply GPS-Health to a wide range of clinical applications such as connecting patients to the right combination of necessary supportive services, or to help geographically allocate resource investments to build access to and improve the utilization of healthcare services. Other applications outside of healthcare could include understanding interlocking mechanisms that lead to worse economic, political, and educational disparities. Community organizations may be able to use GPS-Health to lobby and advocate for better access to SDOH in their own neighborhoods.

## Data Availability

All data produced are available online at https://doi.org/10.5281/zenodo.14422743

